# Bridging Pleiotropic Mechanisms in Leprosy Type-1 Reactions and Neurodegenerative diseases

**DOI:** 10.1101/2025.06.05.25329059

**Authors:** Vinicius M. Fava, Jônatas Perico, Marianna Orlova, Monica Dallmann-Sauer, Yong Zhong Xu, Nguyen Van Thuc, Vu Hong Thai, Andrea F. Belone, Ana Carla P Latini, Erwin Schurr

## Abstract

Leprosy is an infectious disease that affects predominantly the skin and peripheral nervous system. Sudden episodes of hyperinflammation, known as Type 1 Reactions (T1R), are a main contributor to permanent nerve damage in leprosy. The genetic component associated with the neuro-inflammatory phenotype of T1R displays pleiotropic effects with Parkinson’s disease (PD) risk genes. In this study, we further explored the genetic overlap between PD and T1R and expanded the evaluation of pleiotropic effects between T1R and other neurodegenerative disorders. We first replicated the association of PD-risk rare variants in *PRKN* with T1R in Vietnamese leprosy patients (*P* = 0.04; OR = 3.1). Additionally, we discovered large *PRKN* structural variants only in T1R-affected participants when compared to T1R-free leprosy patients. Analysis of 24 PD associated genes revealed compound effects between rare protein-altering variants and T1R in the interacting genes *PRKN/PINK1* (*P* = 2.7^−05^; OR = 4.0) and a combination of rare/low frequency variants in the *LRRK2/GAK* pair (*P* = 6.7^−05^; OR = 0.54). These findings validated a genetic overlap between T1R and PD with two distinct axes, one of shared risk via *PRKN/PINK1* and a second of antagonistic pleiotropic effects via *LRRK2/GAK*. When testing an additional 94 genes associated with neurodegenerative diseases other than PD, we identified variants in the amyotrophic lateral sclerosis (ALS) disease-risk gene *TBK1* associated with T1R (*P* = 0.004; OR = 12.9). All *TBK1* mutations in T1R-affected participants located to the ubiquitin-like TBK1 domain. *PRKN*, *PINK1* and *TBK1* play a critical role in autophagy and in the host immune response to *Mycobacteria*. Our results highlight shared biological processes between leprosy and neurodegenerative diseases, which may indicate candidate drugs for repurposing for a more favorable T1R management.

**AUTHORS SUMMARY:** In this study, we investigated the shared genetic component between hyperinflammatory processes in leprosy and neurodegenerative diseases. A subset of leprosy patients develop sudden episodes of severe inflammation, known as type-1 reactions (T1R), which frequently lead to permanent nerve injury. Here, we aimed to identify key genetic mediators of nerve damage shared with diseases of the central nervous system and T1R, which affects the peripheral nervous system. Previous studies showed Parkinson’s disease (PD) risk genes associated with T1R. To explore this further, we analyzed 24 well-established PD-risk genes in over 800 Vietnamese leprosy patients. We found that rare mutations in *PRKN* and *PINK1* were more frequent in individuals experiencing T1R. These genes are involved in mitochondrial quality control and responses to intracellular pathogens. Inversely, the *LRRK2* and *GAK* PD-risk genes were associated with protection from T1R. This pattern suggested two independent biological processes influencing T1R susceptibility. Our findings highlight a shared genetic component between PD and T1R and suggest that therapies developed for PD may be repurposed for T1R management. Our results underscore the value of cross-disease research to uncover new therapeutic strategies, particularly for neglected diseases with limited treatments or therapies with frequent adverse effects.

## INTRODUCTION

Leprosy is a curable infectious disease caused by *Mycobacterium leprae* that primarily affects the skin and peripheral nervous system (PNS). Macrophages and Schwann cells, glia that form the myelin sheath in the PNS, are the main host cells of *M. leprae* (1). During the course of leprosy, 30% to 50% of patients experience episodes of hyperinflammation named type-1 reactions (T1R) (2). T1R are characterized by pathological cellular immune responses directed against the sites of infection. If not promptly treated, T1R is a major cause of permanent nerve impairment leading to irreversible damage to the neurons of the PNS. The recommended therapeutical intervention for T1R episodes is prolonged use of corticosteroids which carries significant adverse effects. Hence, the identification of prophylactic measures as well as novel and more tolerable therapies for T1R management remains of major public health priority in leprosy endemic areas.

We previously established a genetic component that predisposes leprosy patients to T1R (3–5). A subset of these genetic factors exhibited pleiotropic effects between T1R and Parkinson’s Disease (PD) (6, 7). To deepen our understanding of these pleiotropic mechanisms we employed a stepwise testing approach. First, we focused on the replication of the association of PD risk genes *PRKN* and *LRRK2* with T1R in Vietnamese leprosy patients. Next, we evaluated if pleiotropic effects extended to other well-established PD-risk genes. Finally, we investigated if T1R shares genetic risk factors with other neurodegenerative diseases (e.g., Alzheimer’s disease, amyotrophic lateral sclerosis, dementia, and dystonia).

We replicated the association of rare protein-altering variants in *PRKN* with T1R in Vietnam. Additionally, we identified two distinct axes of pleiotropic effects between T1R and neurodegenerative disease risk genes: one axis of shared risk factors via *PRKN*, *PINK1*, *ADORA1,* and *TBK1* involved in mitochondrial homeostasis, cellular stress, and host response to intracellular bacteria, and a second axis of antagonistic pleiotropy via *LRRK2* and *GAK*.

## RESULTS

### Protein-altering variants of neurodegenerative disease risk genes found among leprosy patients

For this study we assembled 412 T1R-affected and 419 T1R-free Vietnamese leprosy patients matched by sex, age, and distribution of leprosy clinical forms (**Table 1**). Of these, 430 individuals had been evaluated in our previous study reporting pleiotropic effects of *PRKN* and *LRRK2* between T1R and PD, while 403 were newly included participants. For all subjects, we sequenced the complete loci of 118 curated neurodegenerative diseases associated genes included in Illumina’s TruSeq Neurodegeneration Panel. Of these genes, 24 have established association with PD (**Fig. S1**). We then performed variant calling and filtering for protein-altering and structural variants (SVs), which were subsequently tested for association with T1R. In total, we detected 1,223 protein-altering variants across 105 genes, while 13 genes had no protein variants among the study participants (**Table S1**). Twelve of the 1,223 variants were large SVs of which eleven were found in PD-risk genes, including four variants longer than 100 kilobases in *PRKN* (**Table S1** and **Fig 1**).

**Fig 1.**
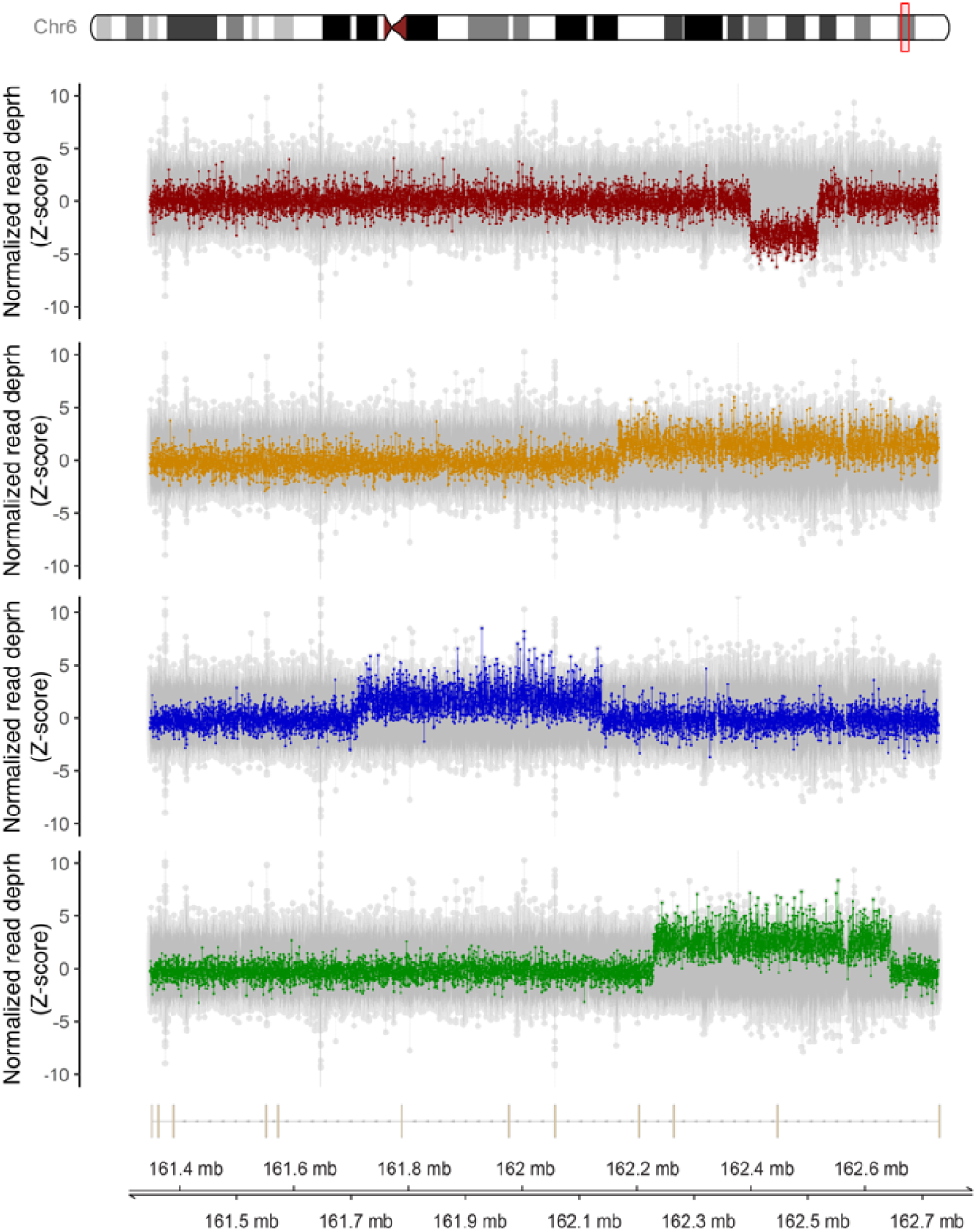
Structural variants in *PRKN* detected in T1R-affected participants. The four panels display the Z-scaled and PCA normalized depth of coverage on the y-axes across the *PRKN* locus shown on the x-axis. Each dot represents the depth of coverage for a probe at a specific chromosomal position within the locus. The coverage for the SV-presenting samples in each plot is highlighted in colors with the remaining 821 samples analyzed for structural variants shown in grey for comparison. *PRKN* structure is shown at the bottom with vertical lines correspond to exons.

**Table 1.** Population phenotypic characteristics.

|  | T1R-free | T1R-affected |
| --- | --- | --- |
| **Sex** |  |  |
| Male | 293 | 295 |
| Female | 119 | 124 |
| **R&J Classification** |  |  |
| BT | 92 | 92 |
| BB | 173 | 176 |
| BL | 148 | 151 |
| **Age** |  |  |
| Mean (SD) | 23.56 (8.1) | 22.83 (9.2) |
| Min – Max | 5 – 67 | 5 – 66 |

BT, borderline tuberculoid; BB, borderline leprosy; BL, borderline lepromatous; R&J, Ridley and Jopling classification of leprosy clinical forms; SD, Standard deviation; T1R, Leprosy Type-1 reaction.

### Replication of pleiotropic effects for PRKN between T1R and PD

We first tested if the burden of rare protein-altering variants (defined as minor allele frequency [MAF] < 1%) in *PRKN* and *LRRK2* differed between the 203 T1R-affected vs 200 T1R-free leprosy patients that had not been included in the previous study. We observed a significant accumulation of protein-altering variants for the *PRKN* gene in the T1R-affected cases as a direct replication of initial findings (**Table 2**). In the new T1R sample set we detected four rare heterozygote SVs, three resulted in multiple *PRKN* exons duplication and one in exon 2 deletion. We also detected one *PRKN* frameshift mutation (T125Rfs). These mutations likely result in pronounced changes on Parkin structure and stability and were all found in T1R-affected cases (**Fig 2**). We also detected a depletion of protein-altering variants for *LRRK2* in T1R-affected compared to T1R-free participants as we reported previously, however, this did not reach statistical significance (**Table 2**).

**Fig 2.**
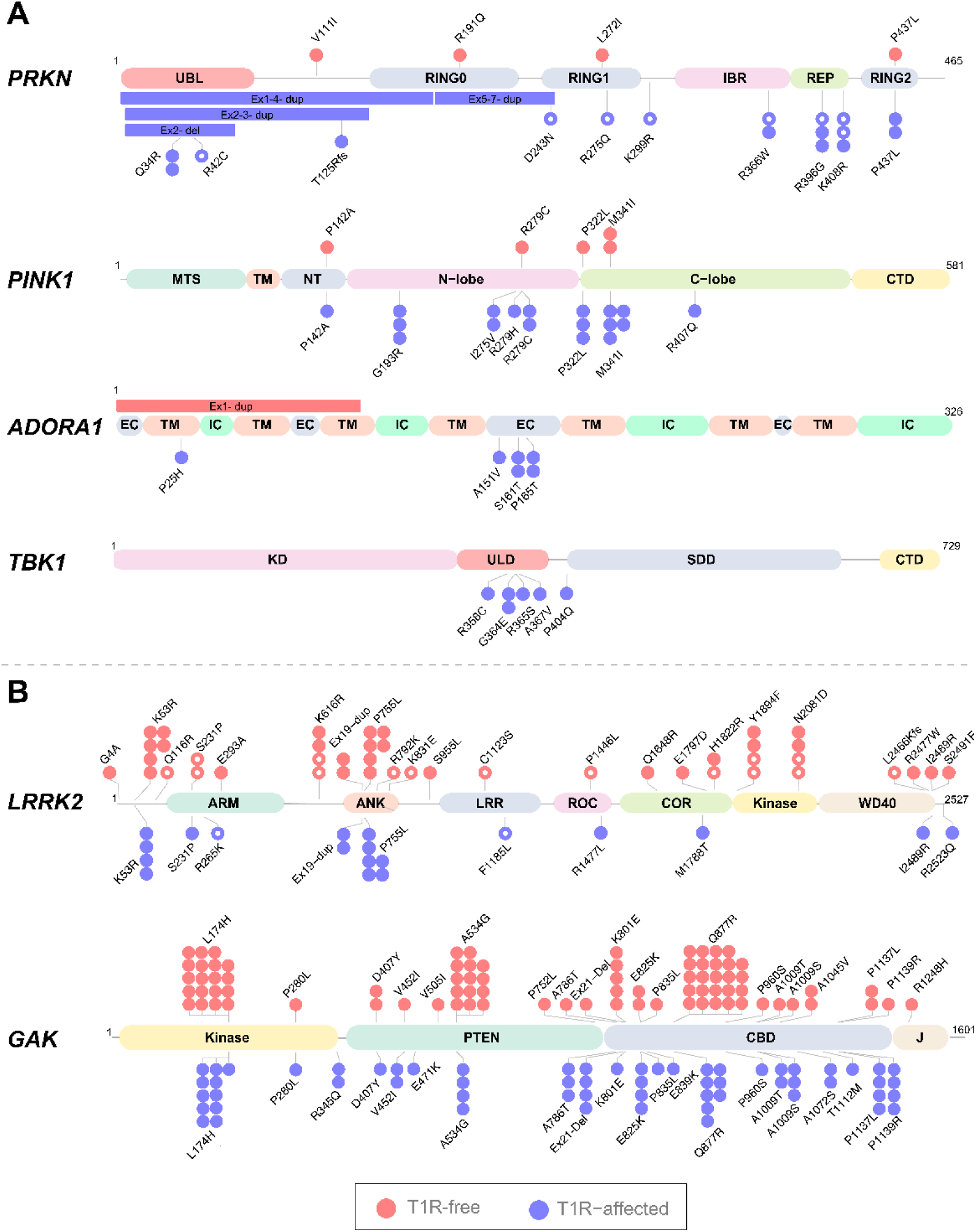
Protein-altering variants in neurodegenerative risk genes associated with T1R. Each dot represents an allele detected in participants with protein-altering variants with minor allele frequency below 1% for each given gene. For *GAK* three low frequency variants associated with T1R protection were also shown in the plot. Each bar represents a SV. Dots and bars were colored according to the T1R status and indicate the protein domain where the variants are located. Alleles detected in our previous study (ref. 6) are indicated with an open circle. (**A**) Rare alleles enriched in T1R-affected and (**B**) rare alleles associated with T1R-protection, i.e. depleted in T1R subjects. UBL, UBiquitin-Like domain; RING Really Interesting New Gene finger domains, IBR, In-Between RING domain; REP, Repressor Element of Parkin; MTS, Mitochondrial Targeting Sequence domain; TM, Transmembrane domain; NT, N-Terminal tail domain; N-lobe and C-lobe of the Kinase domain; CTD, C-Terminal domain; KD, Kinase Domain; ULD, Ubiquitin-Like Domain; SDD, Scaffold Dimerization Domain; EC, Extracellular domain; IC, Intracellular domain. ARM, Armadillo repeats; ANK, Ankyrin repeats; LRR, Leucine-rich Repeats; ROC, Ras of Complex Proteins; COR, C-terminal of ROC. CBD, Clathrin-binding region.

**Table 2.** Replication of accumulation of rare protein-altering variants in the *PRKN* and *LRRK2* genes in T1R patients.

| Gene | T1R-affected | T1R-free | Cases/Controls | *P* value | OR (95% CI) | Samples |
| --- | --- | --- | --- | --- | --- | --- |
| *PRKN* | 1.9% | 0.0% | 216/214 | 0.003 | 7.61 (1.97 - 29.4) | Ref. (6) |
|  | 3.2% | 1.0% | 204/200 | 0.04 | 3.13 (1.04 - 9.37) | New |
| *LRRK2* | 2.8% | 6.3% | 216/214 | 0.01 | 0.43 (0.23 - 0.82) | Ref. (6) |
|  | 2.7% | 4.3% | 204/200 | 0.36 | 0.69 (0.31 - 1.54) | New |

Rare was defined as minor allele frequency < 1%; T1R, Leprosy Type-1 reaction. OR, odds ratio; CI, confidence interval.

### Additional PD-risk genes show pleiotropic effects with leprosy T1R

Next, we combined the 833 participants in a single population set and compared the burden of protein-altering variants between T1R-affected vs T1R-free participants for *PRKN*, *LRRK2*, and 22 additional PD-risk genes included in the neurodegenerative disease panel. We then performed a global analysis to estimate *PRKN* and *LRRK2* effect sizes in T1R susceptibility by combining the sample sets of the previous study with the current one (**Table 3**). Of note, we did not observe within gene compound heterozygotes for *PRKN*. In addition, we tested if T1R-affected cases or T1R-free controls carried different burden of protein-altering variants in 22 additional PD-risk genes that were not assessed in our previous study (**Table S2**). Three additional PD-associated genes (*PINK1, ADORA1* and *GAK*) showed significant differential burden of protein-altering variants between cases and controls at false discovery rate (FDR) < 5%. Rare protein-altering variants in *PINK1* and *ADORA1* were more frequent in T1R-affected cases while for GAK a protection effect was significant only when low frequency variants (MAF < 5%) were included (**Table 3** and **Fig 2**).

**Table 3.** Genewise association of rare variants with T1R in 833 participants.

| Gene | Disease | MAF | Variants<br>(count) | T1R<br>affected | T1R<br>free | *P* value <sup>a</sup> | *FDR*<br>*P* value <sup>b</sup> | OR (95% CI) |
| --- | --- | --- | --- | --- | --- | --- | --- | --- |
| *GAK* | PD | < 1% | 22 | 3.8% | 3.7% | 0.44 | 0.73 | 0.80 (0.45 – 1.42) |
| *GAK* | PD | < 5% | 25 | 5.9% | 8.5% | 0.004 | 0.03 | 0.53 (0.35 – 0.82) |
| *LRRK2* | PD | < 1% | 27 | 2.2% | 5.1% | 0.001 | 0.02 | 0.39 (0.22 – 0.70) |
| *LRRK2* | PD | < 5% | 29 | 5.5% | 9.2% | 0.006 | 0.03 | 0.55 (0.37 – 0.84) |
| *ADORA1* | PD | < 1% | 5 | 0.7% | 0.1% | 0.007 | 0.03 | 10.0 (1.89 – 53.4) |
| *PINK1* | PD | < 1% | 8 | 2.0% | 0.6% | 0.002 | 0.02 | 4.30 (1.68 – 11.0) |
| *PRKN* | PD | < 1% | 17 | 2.4% | 0.5% | 0.003 | 0.02 | 3.77 (1.54 – 9.23) |
| *TBK1* | ALS | < 1% | 5 | 0.7% | 0.0% | 0.004 | 0.05 | 12.9 (2.22 – 75.1) |
| *GAK – LRRK2* | . | < 5% | 54 | 11.4% | 17.7% | 6.7E<sup>-5</sup> | . | 0.54 (0.40 – 0.73) |
| *PRKN – PINK1* | . | < 1% | 25 | 4.4% | 1.1% | 2.7E<sup>-5</sup> | . | 4.01 (2.10 – 7.68) |
| *PRKN – PINK1 –*<br>*TBK1* | . | < 1% | 30 | 5.1% | 1.1% | 8.6E<sup>-7</sup> | . | 4.61 (2.51 – 8.49) |
| *PRKN – PINK1 –*<br>*TBK1 – ADORA1* | . | < 1% | 35 | 5.8% | 1.2% | 2.8E<sup>-8</sup> | . | 5.05 (2.85 – 8.96) |

<sup>a</sup> *P* values estimated with the inverse of variance meta-analysis. <sup>b</sup> Benjamini–Hochberg false discovery rate (FDR) *P*
values. T1R, Leprosy Type-1 reaction. T1R-affected (N= 420). T1R-free (N= 414). OR, odds ratio; CI, confidence interval;
PD, Parkinson's disease; ALS, Amyotrophic lateral sclerosis; MAF, Minor allele frequency.

The proteins encoded by *PINK1* and *PRKN,* as well as those encoded by *LRRK2* and *GAK,* are two interaction pairs. Hence, we hypothesized that mutations in the corresponding gene pairs may have additive effects by affecting the same molecular signalling pathways. We tested if the combined mutation burden in each pair differed significantly between T1R-affected and T1R-free subjects. We observed a strong association of rare protein-altering variants in *PRKN* – *PINK1* with T1R-affected (**Table 3**). The burden of protein-altering variants in *GAK* – *LRRK2,* which was associated with protection from T1R, was primarily driven by independent variants (LD *r*^2^ = 0) within the 1% – 5% MAF range. These included three nonsynonymous variants in *GAK* (L174H, A534G, and Q877R), and the R1628P mutation in *LRRK2* (**Table 4**). Notably, the association of *LRRK2* with T1R-free remained significant even in the absence of the low frequency R1628P mutation (**Table 3**). *LRRK2* mutations with MAF < 1% were more frequent in the critical ROC/COR and Kinase domains among T1R-free participants (**Fig. 2**). Taken together, our results confirmed and expanded the existence of two axes of opposing pleiotropic effects between T1R and PD: the *PRKN/PINK1* axis, which predisposes to T1R, and the *LRRK2/GAK* axis, which confers T1R protection while both axes confer risk to PD.

**Table 4.** Low frequency protein-altering variants in *GAK* and *LRRK2*.

| Gene | Variants<sup>a</sup> | T1R<br>affected | T1R<br>free | N | *P*<br>value | OR (95% CI) | CADD<br>Score | gnomAD |  |
| --- | --- | --- | --- | --- | --- | --- | --- | --- | --- |
|  |  |  |  |  |  |  |  | Global | EAS |
| *GAK* | Q877R | 0.96% | 2.91% | 416/413 | 0.009 | 0.36 (0.16 – 0.77) | 3.99 | 0.03% | 0.77% |
|  | A534G | 0.51% | 1.79% | 390/390 | 0.004 | 0.22 (0.08 – 0.62) | 25.2 | 0.003% | 0.08% |
|  | L174H | 1.34% | 2.31% | 410/411 | 0.02 | 0.40 (0.18 – 0.88) | 26.8 | 0.003% | 0.01% |
| *LRRK2* | R1628P | 2.80% | 4.09% | 393/379 | 0.12 | 0.63 (0.34 – 1.13) | 27.5 | 0.09% | 0.84% |
|  | G2385R | 1.20% | 0.97% | 374/361 | 0.54 | 1.39 (0.49 – 3.90) | 23.2 | 0.08% | 2.66% |

<sup>a</sup> Linkage disequilibrium (LD) $r^2 = 0$ for the three *GAK* and the two *LRRK2* variants. T1R, leprosy Type-1 reaction; N, number of T1R-affected by T1R-free participants; OR, odds ratio; CI, confidence interval; CADD, Combined Annotation-Dependent Depletion; EAS, East Asian populations of gnomAD.

### Pleotropic effects for T1R and neurodegenerative diseases risk genes

In addition to PD, the TruSeq Neurodegeneration Panel includes 94 genes curated as either causal or strong risk factors for Alzheimer’s disease (ALZ), amyotrophic lateral sclerosis (ALS), frontotemporal dementia (FTD), dystonia, and other rare neurodegenerative conditions such as Huntington’s disease and Cerebellar Ataxia (**Fig S1**). We tested if any of these genes were associated with T1R (**Table S2**). We found protein-altering variants in the ALS-risk gene *TBK1* associated with susceptibility to T1R at FDR of 5% (**Table 3**). All *TBK1* mutation (N = 5) were observed in T1R-affected cases and located near or in the ubiquitin-like domain of the protein (**Fig 2**). No other gene surpassed the suggestive FDR < 10% in the studied population (**Table S2**).

### Genes of mitochondrial checkpoint are risk factors for T1R and neurodegenerative diseases

Mutations in *TBK1* are most commonly associated with ALS and FTD. While parkinsonism syndrome is often observed in FTD cases, there is no substantial evidence that *TBK1* mutations are associated with PD. However, *TBK1,* together with *PRKN* and *PINK1,* plays a critical role in mitochondrial quality control. Therefore, we hypothesized that genes involved in mitochondrial checkpoints represent a biological mechanism underlying pleiotropy between T1R and neurodegenerative diseases. We performed a meta-analysis combining the effects of *PRKN*, *PINK1*, and *TBK1*, and observed a more significant association with T1R (**Table 3**). We then expanded the meta-analysis to include *ADORA1*, a regulator of cellular stress that contributes to mitochondrial stability. The combined analysis of these four genes revealed a high effect size, suggesting a substantial contribution to T1R susceptibility in the Vietnamese population (*P* = 2.8E^−8^; OR = 5.05; confidence interval 95% (2.85 – 8.96)).

## DISCUSSION

We identified two distinct axes of pleiotropic effects between leprosy T1R and PD: one involving *LRRK2* and *GAK,* and another comprising the *PRKN* and *PINK1*. *LRRK2* and *GAK* are associated with both sporadic and familial forms of late-onset PD (LOPD) (8). Nonsynonymous variants in *LRRK2* resulting in amino acid substitutions at positions 2019 and 1441 in the ROC/COR and kinase domains are known to cause LOPD, while regulatory variants of *GAK* have been shown to modulate PD risk across multiple ethnicities (9, 10). Conversely, autosomal recessive mutations in *PRKN* and *PINK1* are the two most prevalent causes of early-onset PD (EOPD). The differences in age of onset and inheritance patterns between these two axes suggest that they may influence distinct aspects of PD pathogenesis. Our findings support the existence of two distinct mechanisms as LOPD-risk genes were associated with T1R-protection (i.e., antagonistic pleiotropy), whereas the EOPD-risk axis showed pleiotropy for both PD and T1R.

In the antagonistic axis, LRRK2 is a multifunctional kinase that regulates various cellular processes, while GAK plays a crucial role in clathrin-mediated vesicle trafficking (10). LRRK2 and GAK function as interaction partners in a complex facilitated by BCL2-associated athanogene 5 (BAG5) (11). BAG5-mediated interaction occurs via the COR and kinase domains of LRRK2, the regions where the mutation burden differed most significantly between T1R-free and T1R-affected individuals, and via the PTEN domain of GAK, which harbors the T1R-protective variant A534G. The LRRK2/GAK complex promotes clearance of Golgi-derived vesicles through autophagy (11). In the context of PD, defects in vesicle trafficking and clearance are hallmarks of α-synuclein accumulation in the brain. In leprosy and T1R, mycobacteria use the host’s autophagy machinery to evade the immune system and survive. However, the mechanism by which LRRK2 variants modulate this process deserves further investigation. For example, LRRK2/GAK mutations could result in gain of function, as we have demonstrated for reactive oxygen species (ROS) production in the LRRK2 R1628P variant (6), which may facilitate vesicle clearance during the early stages of infection and prevent hyperactivation of the host immune system.

With respect to risk pleiotropic effects, we replicated our prior findings of rare protein-altering variants in *PRKN* as T1R risk factors (6). In addition, we detected the same large SVs observed in cases of EOPD in T1R-affected participants (12). Homozygous SVs or compound heterozygous SVs in combination with *PRKN* protein-altering variants represent up to 72% of *PRKN* mutations in Caucasian and Chinese EOPD cases (12–15). In EOPD, SVs occur most frequently in *PRKN* exons 2, 3, and 7, similar to what we observed in T1R-affected cases. However, in the heterozygous state and in the absence of additional mutations in other EOPD-risk genes, *PRKN* SVs are not associated with an increased risk of PD (15, 16). This difference in dominance effect between T1R and PD is consistent with the lack of epidemiological evidence suggesting a higher incidence of PD in T1R or leprosy affected individuals. Despite this, healthy carriers of *PRKN* heterozygote mutations have shown increased serum levels of pro-inflammatory cytokines (e.g., IL-6, IL-1β, CCL2) similar to those observed in homozygote *PRKN* mutation carriers with PD (17). This suggests that *PRKN* modulates the extent of the immune response irrespective of the T1R or PD phenotype. Moreover, in a mouse model the pro-inflammatory state associated with loss of *PRKN* activity was STING-mediated (17). This provides a direct link to *TBK1*, a critical downstream effector in STING signaling (17). In our study, we only observed mutations in the ubiquitin-like domain of *TBK1* and only in T1R participants. STING –TBK1 play a pivotal role in autophagy, antimicrobial defence, and antigen presentation (18, 19). Deletion of the UBL domain was shown to impair TBK1 kinase activity (20) which plays a key role in autophagosome formation and pro-inflammatory signalling (18, 19). The key role of *PRKN* signalling in both T1R and PD was further highlighted by our finding of an enrichment of *PINK1* mutations in T1R patients. The most frequent *PINK1* mutations associated with EOPD are nonsynonymous variants located in the kinase domain. A similar pattern was observed in T1R, where the vast majority of *PINK1* mutations clustered in the C- and N-lobe of the PINK1 kinase domain. Phosphorylation of Parkin by PINK1 is required to activate its autoinhibited Parkin ubiquitin ligase domain (21). The functional role of *PRKN/PINK1* activity is among the best characterized mechanisms in the pathogenesis of EOPD.

The interplay between PINK1 and Parkin is crucial for selective autophagy and immune response processes, such as mitophagy, mitochondrial antigen presentation via mitochondrial-derived vesicles (MitAP), and regulation of adaptive immunity. These biological processes may represent the key components underlying the pleiotropic effect between T1R and PD. In mitophagy, PINK1 and Parkin mark damaged mitochondria for degradation through ubiquitination. Failure to remove damaged mitochondria leads to oxidative stress via increase in intracellular production of ROS (22). Stimulation of adenosine receptors, including the T1R-risk gene *ADORA1*, protect against oxidative damage during neuroinflammation (23, 24). Therefore, defects in mitochondrial homeostasis and cellular stress might be linked to inflammation in both PD and T1R. In xenophagy, a host response mechanism against intracellular pathogens, PINK1 and Parkin ubiquitinate phagosomes containing intracellular pathogens. However, certain bacteria, such as *M. tuberculosis*, escape xenophagy, survive and replicate in *PRKN* knockout macrophages (*25, 26*). Defects in PINK1/Parkin-mediated mycobacterial clearance could explain the previously observed association between *PINK1* and *PRKN* with leprosy *per se* (27–29). In T1R, the contribution of xenophagy had dichotomous findings with T1R episodes linked to blockade and activation (30). Therefore, alternative mechanisms such as MitAP are candidates for the underlying biological role of *PRKN/PINK1* in T1R. In MitAP, PINK1 and Parkin suppress mitochondrial antigen presentation to immune cells (31). Loss of *PRKN* and *PINK1* function impairs this suppression, which could contribute to the immune mediated neurodegeneration in PD and the excessive inflammation observed in T1R (17). In PD, MitAP functions independently of autophagy; however, the extent to which this applies to T1R remains unknown.

The dynamics of the immune response are controlled by a complex interplay between genomic and environmental factors, where individual genes often regulate multiple aspects of disease pathogenesis. To illustrate this, excessive inflammation increases the risk of tissue damage while insufficient inflammation predisposes a person to infections or facilitates pathogen dissemination. Thus, maintaining a balance between protective and damaging responses is essential for immunological homeostasis. A compelling example is the *LRRK2* R1628P mutation, which has been associated with T1R-protection when comparing T1R-affected and T1R-free leprosy patients (6). However, when each T1R subgroup (affected and unaffected) was compared independently to healthy controls, the R1628P mutation showed an opposing effect (32). The 1628P allele was protective in the T1R-affected subgroup but associated with susceptibility to leprosy *per se* in the T1R-free group. Interestingly, the association of R1628P with leprosy *per se* was also observed in the Chinese population (33). This seemingly paradoxical observation exemplifies how a single variant can shape the outcome of leprosy and its endophenotypes.

The identification of pleiotropic biological processes offers valuable opportunities for therapeutic repurposing. This strategy is particularly important for neglected tropical diseases such as leprosy and T1R, where research and development resources are limited. In contrast, several drugs and compounds targeting PD-risk genes including *PRKN*, *PINK1*, GAK and *LRRK2* are currently under active investigation or enrolled in clinical trials for PD (34, 35). Although standard PD treatments such as dopamine-targeting drugs (e.g., levodopa), are unlikely to be relevant for leprosy or T1R, compounds that modulate pleiotropic genes may be promising candidates for repurposing in leprosy/T1R. For example, the allosteric modulator tetrahydropyrazolo-pyrazine (THPP) functions as a molecular glue that enhances the activity of Parkin’s UBL domain (36). THPP has been shown to partially rescue Parkin UBL activity in EOPD cases carrying R42P mutation, the same residue observed mutated in T1R-affected subjects affected by SVs. However, the rescue effect of THPP is limited to the ubiquitin function of Parkin, which may constrain its therapeutic relevance for T1R where additional mutations in different domains may impair other Parkin functions or interactions.

Another promising candidate for repurposing is the small molecule Kinetin riboside, which stimulates PINK1-dependent mitophagy (37, 38). In parallel, additional patents have been filed for small molecules also targeting PINK1 and Parkin-mediated mitophagy (39). However, several of these molecules showed limited effects in PD rodent models and will requiring further optimization and development (19, 34). An alternative strategy currently being explored in PD involves inhibiting Parkin deubiquitinating enzymes (40). For instance, a USP30 knockout has been shown to rescue mitophagy defects caused by pathogenic Parkin mutations. Compounds targeting ubiquitin-specific proteases, including USP30, and other deubiquitinases such as ataxin-3 are under clinical investigation (41, 42). Among the most advanced candidates are substituted cyanopyrrolidines (40). Molecules targeting the T1R antagonistic pleiotropic genes *LRRK2* and *GAK* are also under investigation in PD (34). However, due to the mutation-dependent effects of LRRK2 in leprosy and its associated endophenotypes, a more tailored selection of candidate compound will be necessary. While many of these compounds are not yet clinically available, partnerships between academic researchers and the commercial developers could provide valuable insights into their broader application. Such collaborations may facilitate the repurposing of both existing and novel therapies for leprosy patients with T1R, who currently rely heavily on long-term corticosteroid treatments.

In conclusion, we identified shared biological processes involving excessive inflammatory responses in leprosy with known PD-risk genes. Our findings convey two important messages: First, they reinforce the hypothesis of an inflammatory component in PD, potentially mediated by an infectious agent, as proposed in a brain-gut axis model. Second, they highlight that excessive inflammation with resulting neuronal damage, in the PNS for leprosy and CNS for PD, is mediated by a shared set of genes that serve as key checkpoints in immune-mediated processes.

## MATHERIALS AND METHODS

### Population sample and targeted sequencing of neurodegenerative diseases risk genes

Written informed consent was obtained from all participants or from parent/guardian of minors. The study was approved by the Research Ethics Board at the Research Institute at McGill University Health Centre in Montreal (REC98-041) and the regulatory authorities of Ho Chi Minh City in Vietnam (So3813/UB-VX and 4933/UBND-VX). We retrieved DNA from 940 leprosy cases from our biobanked samples of the Vietnamese population. Of these, 470 were leprosy cases presenting at least one T1R episode, and 470 were T1R-free leprosy patients absent of any type of leprosy reactions matched by sex, age of onset and clinical form of the disease. A DNA aliquot of 50 ng/uL from each participant was then used to target sequence PD-risk genes with the TruSeq Neurodegeneration Panel. This panel was designed by Illumina with input of neurodegenerative researchers to target sequence the entire locus of 118 selected genes (∼8.6 mega bases). In addition to Parkinson’s disease risk genes, the panel covers a broad spectrum of neurodegenerative conditions such as Alzheimer’s disease, ALS, Frontotemporal Dementia among other rare neurodegenerative disorders (**Fig S1**). Libraries were prepared according to Illumina manufacture instructions using 50 ng/uL DNA as input. TrueSeq libraries from all participants were 150bp paired-end sequenced with Illumina HiSeq 4000.

### Variant calling and quality control

Sequenced libraries were processed in the high-performance computational platform Cedar from the Digital Research Alliance of Canada. Nextera adaptors were trimmed using TrimGalore v0.4.5. Reads were aligned to the human genome hg38 build using BWA mem v.0.7.17 with default parameters and duplicate reads were marked with PICARD v.2.18.9. Subjects with mean depth of coverage < 10X or samples with relatedness > 0.1 were excluded from further analysis. Of the 940 samples 833 passed quality control. For these participants we obtained a mean depth of coverage of 20X per gene in the Neurodegenerative panel (**Fig S2**). Next, variants were called using HaplotypeCaller from GATK v.4.0.8.1 and DeepVariant v.1.0 in WES mode. Cohort genotyping of individual gVCFs was performed with GenotypeGVCFs from GATK and GLNexus for DeepVariant. Variants with Q < 30, Depth < 10X, and missing > 20% were excluded. Protein-altering variants with consensus calls in the two approaches were then kept for analysis. Singletons identified with less than 15 reads in the associated genes were validated by deep sequencing with a custom panel from Twist Bioscience and processed with the same pipeline.

### Structural variants detection

The probes designed for the neurodegeneration panel captured continuous regions of the genome, including gene flanking regions, exons, introns, and promoters of the targeted genes. Hence, the depth of coverage of sequential probes can be used to estimate the occurrence of SVs. We used the probe location to estimate the depth of coverage by probe per sample to detect large SVs such as deletions, duplications and copy number variants (CNVs). Briefly, reads with Phred > 20 were used to estimate the depth per probe loci using GATK *DepthOfCoverage* for the 833 participants. We then used GATK *GCContentByInterval* to detect regions with extreme GC content and plinkseq v.0.10 to detect regions of low complexity. These regions could interfere with data normalization and were excluded from the SV calling. Next, XHMM was used to perform a principal component (PC) analysis with the normalized counted data. We then regressed out the top 19 principal components that cumulatively explained 70% of the total variance and Z-scaled the data per sample as per recommendation. To detect SVs, XHMM --discover and --genotype was used with default parameters and distance between targets of 500bp. As quality control filtering, SVs with > 15 sequential probes deviating one standard deviation from the population mean Z-scaled coverage and encompassed exons were selected for analysis.

### Data analysis

The stepwise protein-altering variant analysis was performed with regenie v.3.6. First, ∼19 thousand variants (coding and non-coding) with MAF > 1% detected in the 118 targeted genes was used to calculate a null model with --step 1 and --bt to define a binary phenotype. This first step was carried out to capture the genetic contributions to the phenotype while accounting for population structure and relatedness. No covariates were included in the model, as per design T1R-affect cases were matched to T1R-free controls (**Table 1**). Next, regenie --step 2 was used to estimate differences in rare variant burden between cases and controls under additive models or kernel models with --vc-tests skato-acat. The analysis used the prediction file generated in --step 1, a set list including 1,223 protein-altering variants split by their corresponding gene, and --rgc-gene-p to estimate one *P* value per gene in two MAF thresholds of <1% and <5%.

We carried out protein-altering variants testing in three stages: (i) split by the population described in Ref (6), and the newly included participants for the *PRKN* and *LRRK2* genes. We then compared different burden between cases – controls for (ii) PD-risk genes and (iii) genes associated with other neurodegenerative conditions. Genes with < 5 protein-altering variants were excluded from testing. Benjamini–Hochberg FDR corrected *P* values were calculated per disease (e.g, T1R vs PD, T1R vs ALS) to account for multiple testing. To test the combined effect of the interactive proteins we performed an inverse of the variance meta-analysis using β and standard errors of the burden for each gene in R environment. The test of individual low frequency variants for *LRRK2* and *GAK* was carried out in a similar fashion as the genewise tests but without defining a set of variants per gene.

## Data Availability

All data produced in the present work are contained in the manuscript.

## DATA AND CODE AVAILABILITY STATEMENT

NGS data are not publicly available due to participant consent restrictions. NGS data are available for leprosy-related studies by requests to the Bioinformatics Platform at the Research Institute of the McGill University Health Centre. Approval from the applicant’s ethics committee is required for additional work on the data. No personal identification of the studied family will be shared due to consent restrictions. All relevant data are within the manuscript and its Supporting Information files. There were no new pipelines or packages generated in this study. The data analysis were performed using publicly available software as described in methods.

## ACKNOWLEGMENTS

We thank all leprosy patients who participated in this study and members of the ES laboratory for many helpful discussions. We thank the members of the leprosy control program at the Hospital for Dermato-Venereology in Ho Chi Minh City and the staff of the leprosy control program in Southern Việt Nam. We would also like to thank all members of the Desjardins ASAP team for insightful feedback throughout the course of this study. This work was funded by grants from the Leprosy Research Initiative (LRI) FP_22/9 to ES and VMF and by the joint efforts of The Michael J. Fox Foundationfor Parkinson’s Research (MJFF) and the Aligning Science Across Parkinson’s (ASAP) initiative. MJFF administers the grant ASAP 000525 on behalf of ASAP and itself. This research was supported through resource allocation in Cedar high performance computing by Digital Research Alliance of Canada.

## Author contributions

Conceptualization, VMF, ES. Population recruitment, ES, MO, NVT, VHT, AFB, and ACPL. Data Curation, VMF, JP, MO, MD-S and, YZX. Formal Analysis, VMF and JP. Funding Acquisition, VMF, ES and ACPL. Writing of the Original Draft, VMF and ES. Review & Editing, all authors reviewed and contributed to edits of the final manuscript.

## SUPPLEMENT

**Fig S1.**
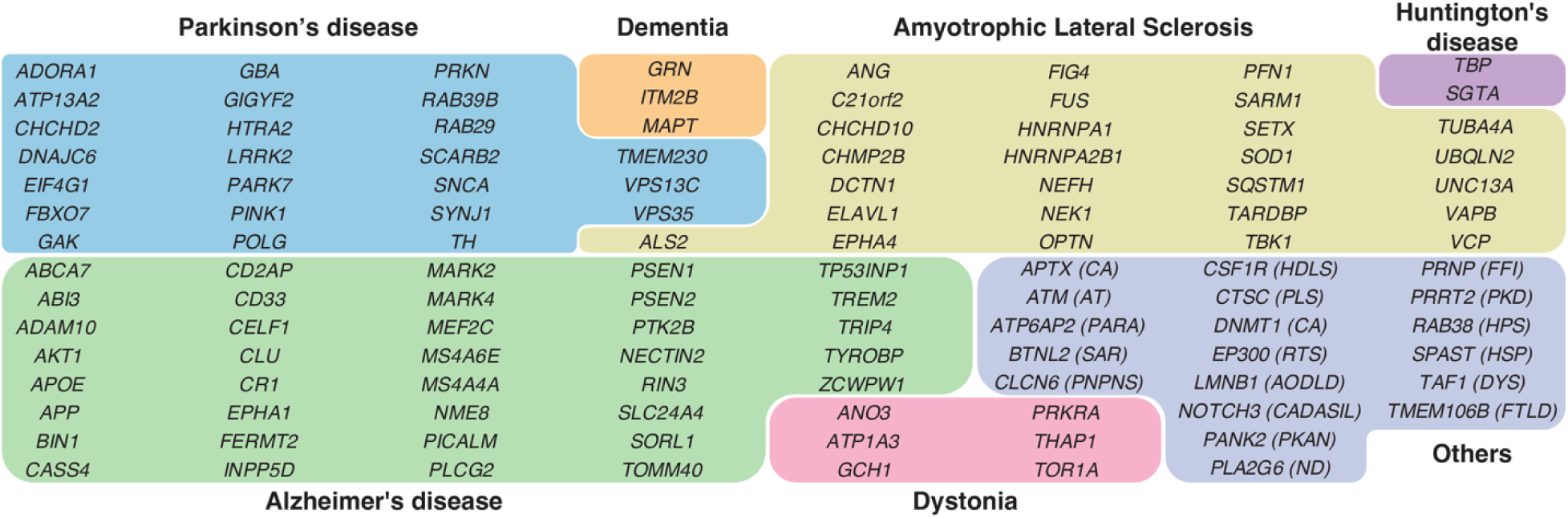
Most frequent phenotype associated with genes in the Neurodegeneration panel. The 118 genes sequenced in the Illumina TruSeq Neurodegeneration Panel^®^ were clustered according to the most frequent phenotype impacted or associated with the gene. The “Others” category included: AODLD, Adult-Onset Leukodystrophy; AT, Ataxia Telangiectasia; CA, Cerebellar Ataxia; CADASIL, Cerebral Autosomal Dominant Arteriopathy with Subcortical Infarcts and Leukoencephalopathy; PNPNS, Childhood-onset progressive neurodegeneration-peripheral neuropathy syndrome; FFI, Fatal familial insomnia; FTLD, Frontotemporal lobar degeneration; HDLS, Hereditary diffuse leukoencephalopathy with spheroids; HSP, Hereditary Spastic Paraplegia; HPS, Hermansky-Pudlak syndrome; ND, Neuroaxonal dystrophy; PKAN, Pantothenate Kinase-Associated Neurodegeneration; PLS, Papillon-Lefèvre Syndrome; PKD, Paroxysmal Kinesigenic Dyskinesia; RTS, Rubinstein-Taybi syndrome; SAR, sarcoidosis; DYS, X-Linked Dystonia; PARA, X-Linked Paraganglioma.

**Fig S2.**
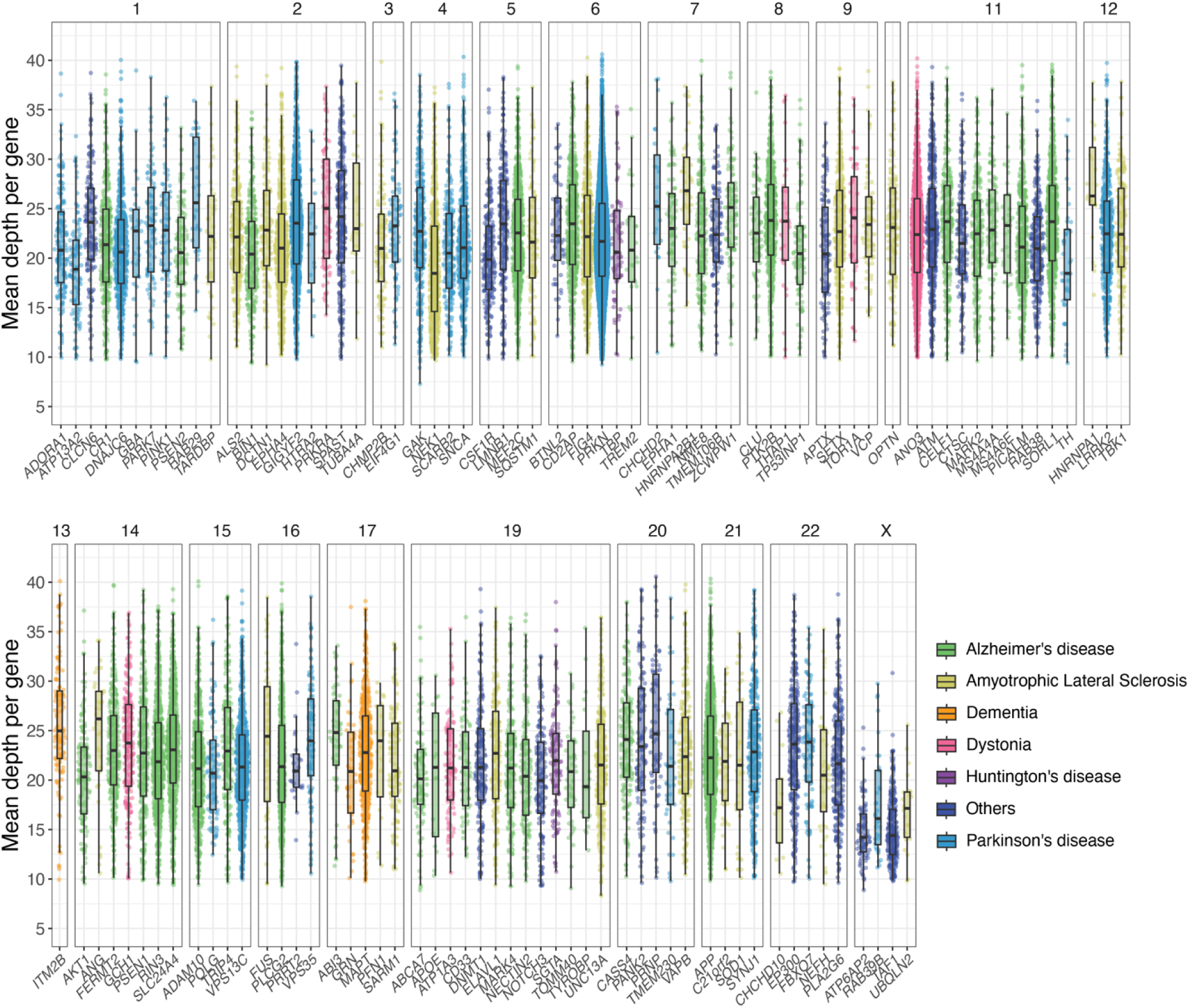
Mean sequencing coverage per gene of the Neurodegeneration panel. Depth of coverage for genes in the Neurodegeneration panel among 876 leprosy-affected subjects. Each dot represents a probe capturing up to 300 base-pairs at the gene loci colored according to the most frequently associated neurological phenotype. The y-axis indicates the mean depth for each gene and the x-axis shows the respective gene (split by chromosome).

**Table S2.**
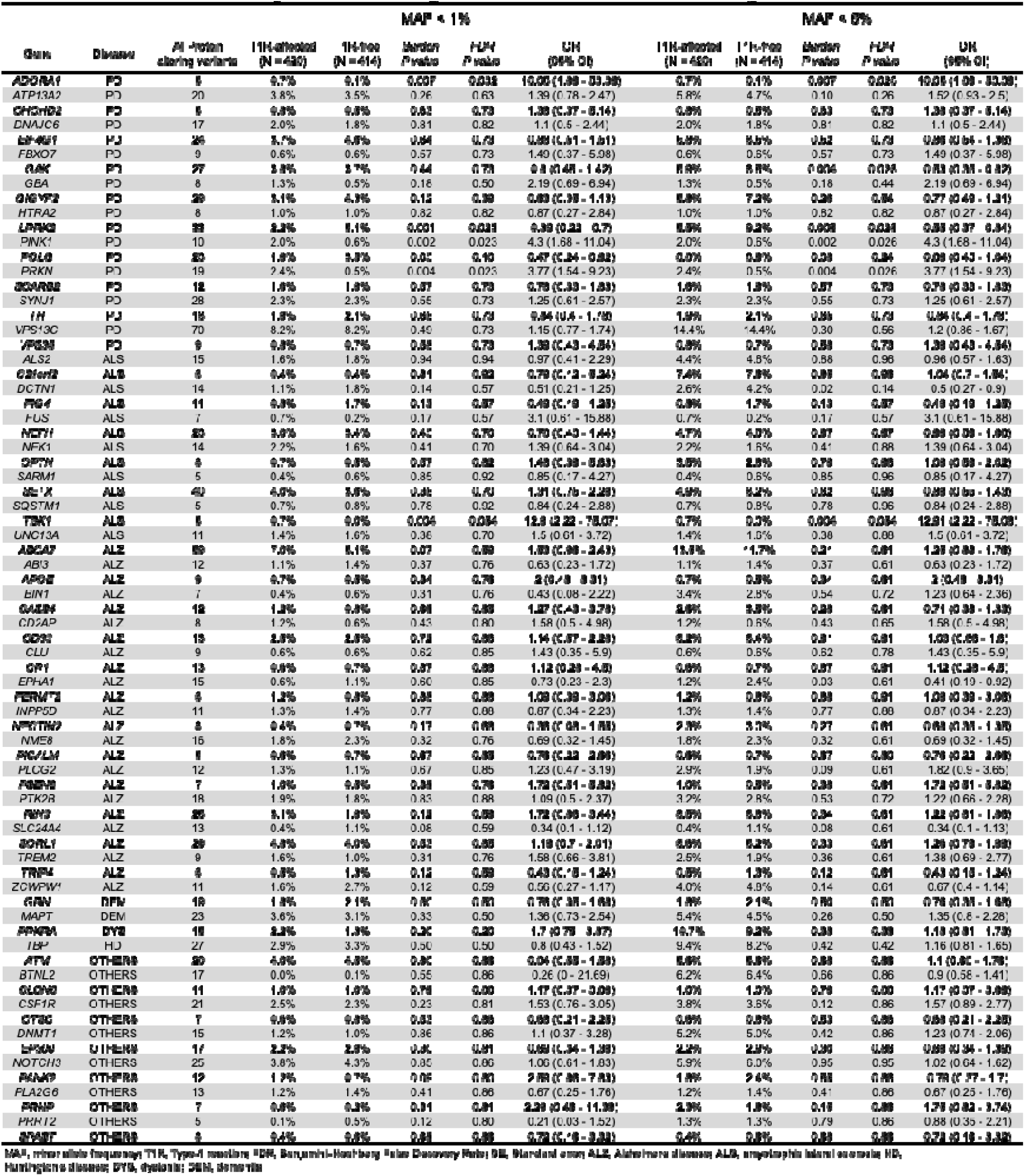
Rare variant analysis in neurodegenerative diseases associated genes.

**Table S1. Protein-altering variants identified in neurodegeneration-associated genes in Vietnamese individuals affected by leprosy.**

## REFERENCES

1. Cabral N, de Figueiredo V, Gandini M, de Souza CF, Medeiros RA, Lery LMS, et al. Modulation of the Response to Mycobacterium leprae and Pathogenesis of Leprosy. Frontiers in microbiology. 2022;13:918009.

2. Fava VM, Dallmann-Sauer M, Schurr E. Genetics of leprosy: today and beyond. Hum Genet. 2020;139(6-7):835–46.

3. Fava VM, Sales-Marques C, Alcais A, Moraes MO, Schurr E. Age-Dependent Association of TNFSF15/TNFSF8 Variants and Leprosy Type 1 Reaction. Front Immunol. 2017;8:155.

4. Fava VM, Manry J, Cobat A, Orlova M, Van Thuc N, Moraes MO, et al. A genome wide association study identifies a lncRna as risk factor for pathological inflammatory responses in leprosy. PLoS Genet. 2017;13(2):e1006637.

5. Fava VM, Cobat A, Van Thuc N, Latini AC, Stefani MM, Belone AF, et al. Association of TNFSF8 regulatory variants with excessive inflammatory responses but not leprosy per se. J Infect Dis. 2015;211(6):968–77.

6. Fava VM, Xu YZ, Lettre G, Van Thuc N, Orlova M, Thai VH, et al. Pleiotropic effects for Parkin and LRRK2 in leprosy type-1 reactions and Parkinson’s disease. Proc Natl Acad Sci U S A. 2019;116(31):15616–24.

7. Fava VM, Manry J, Cobat A, Orlova M, Van Thuc N, Ba NN, et al. A Missense LRRK2 Variant Is a Risk Factor for Excessive Inflammatory Responses in Leprosy. PLoS Negl Trop Dis. 2016;10(2):e0004412.

8. Gilks WP, Abou-Sleiman PM, Gandhi S, Jain S, Singleton A, Lees AJ, et al. A common LRRK2 mutation in idiopathic Parkinson’s disease. Lancet. 2005;365(9457):415-6.

9. Li NN, Chang XL, Mao XY, Zhang JH, Zhao DM, Tan EK, et al. GWAS-linked GAK locus in Parkinson’s disease in Han Chinese and meta-analysis. Hum Genet. 2012;131(7):1089–93.

10. Pankratz N, Wilk JB, Latourelle JC, DeStefano AL, Halter C, Pugh EW, et al. Genomewide association study for susceptibility genes contributing to familial Parkinson disease. Hum Genet. 2009;124(6):593–605.

11. Beilina A, Rudenko IN, Kaganovich A, Civiero L, Chau H, Kalia SK, et al. Unbiased screen for interactors of leucine-rich repeat kinase 2 supports a common pathway for sporadic and familial Parkinson disease. Proc Natl Acad Sci U S A. 2014;111(7):2626–31.

12. Menon PJ, Sambin S, Criniere-Boizet B, Courtin T, Tesson C, Casse F, et al. Genotype-phenotype correlation in PRKN-associated Parkinson’s disease. NPJ Parkinsons Dis. 2024;10(1):72.

13. Towns C, Fang ZH, Tan MMX, Jasaityte S, Schmaderer TM, Stafford EJ, et al. Parkinson’s families project: a UK-wide study of early onset and familial Parkinson’s disease. NPJ Parkinsons Dis. 2024;10(1):188.

14. Hua P, Zhao Y, Zeng Q, Li L, Ren J, Guo J, et al. Genetic Analysis of Patients With Early-Onset Parkinson’s Disease in Eastern China. Front Aging Neurosci. 2022;14:849462.

15. Zhu W, Huang X, Yoon E, Bandres-Ciga S, Blauwendraat C, Billingsley KJ, et al. Heterozygous PRKN mutations are common but do not increase the risk of Parkinson’s disease. Brain. 2022;145(6):2077–91.

16. Hu J, Waters CH, Spiegelman D, Fon EA, Yu E, Asayesh F, et al. Gene-based burden analysis of damaging private variants in PRKN, PARK7 and PINK1 in Parkinson’s disease cohorts of European descent. Neurobiol Aging. 2022;119:136–8.

17. Sliter DA, Martinez J, Hao L, Chen X, Sun N, Fischer TD, et al. Parkin and PINK1 mitigate STING-induced inflammation. Nature. 2018;561(7722):258–62.

18. Pilli M, Arko-Mensah J, Ponpuak M, Roberts E, Master S, Mandell MA, et al. TBK-1 promotes autophagy-mediated antimicrobial defense by controlling autophagosome maturation. Immunity. 2012;37(2):223–34.

19. O’Callaghan B, Hardy J, Plun-Favreau H. PINK1: From Parkinson’s disease to mitophagy and back again. PLoS Biol. 2023;21(6):e3002196.

20. Oakes JA, Davies MC, Collins MO. TBK1: a new player in ALS linking autophagy and neuroinflammation. Mol Brain. 2017;10(1):5.

21. Gladkova C, Maslen SL, Skehel JM, Komander D. Mechanism of parkin activation by PINK1. Nature. 2018;559(7714):410–4.

22. Picca A, Calvani R, Coelho-Junior HJ, Landi F, Bernabei R, Marzetti E. Mitochondrial Dysfunction, Oxidative Stress, and Neuroinflammation: Intertwined Roads to Neurodegeneration. Antioxidants (Basel). 2020;9(8).

23. Mangmool S, Kyaw ETH, Nuamnaichati N, Pandey S, Parichatikanond W. Stimulation of adenosine A(1) receptor prevents oxidative injury in H9c2 cardiomyoblasts: Role of Gbetagamma-mediated Akt and ERK1/2 signaling. Toxicol Appl Pharmacol. 2022;451:116175.

24. Marti Navia A, Dal Ben D, Lambertucci C, Spinaci A, Volpini R, Marques-Morgado I, et al. Adenosine Receptors as Neuroinflammation Modulators: Role of A(1) Agonists and A(2A) Antagonists. Cells. 2020;9(7).

25. Manzanillo PS, Ayres JS, Watson RO, Collins AC, Souza G, Rae CS, et al. The ubiquitin ligase parkin mediates resistance to intracellular pathogens. Nature. 2013;501(7468):512–6.

26. Behr MA, Schurr E. Cell biology: A table for two. Nature. 2013;501(7468):498–9.

27. Alter A, Fava VM, Huong NT, Singh M, Orlova M, Van Thuc N, et al. Linkage disequilibrium pattern and age-at-diagnosis are critical for replicating genetic associations across ethnic groups in leprosy. Hum Genet. 2013;132(1):107–16.

28. Mira MT, Alcais A, Nguyen VT, Moraes MO, Di Flumeri C, Vu HT, et al. Susceptibility to leprosy is associated with PARK2 and PACRG. Nature. 2004;427(6975):636–40.

29. Wang D, Zhang DF, Feng JQ, Li GD, Li XA, Yu XF, et al. Common variants in the PARL and PINK1 genes increase the risk to leprosy in Han Chinese from South China. Sci Rep. 2016;6:37086.

30. Cerqueira DDN, Pereira ALS, da Costa AEC, de Souza TJ, de Sousa Fernandes MS, Souto FO, et al. Xenophagy as a Strategy for Mycobacterium leprae Elimination during Type 1 or Type 2 Leprosy Reactions: A Systematic Review. Pathogens. 2023;12(12).

31. Matheoud D, Sugiura A, Bellemare-Pelletier A, Laplante A, Rondeau C, Chemali M, et al. Parkinson’s Disease-Related Proteins PINK1 and Parkin Repress Mitochondrial Antigen Presentation. Cell. 2016;166(2):314–27.

32. Fava VM, Cobat A, Gzara C, Alcais A, Abel L, Schurr E. Reply to Zhang, et al.: The differential role of LRRK2 variants in nested leprosy phenotypes. Proc Natl Acad Sci U S A. 2020;117(19):10124–5.

33. Zhang DF, Wang D, Yu YL, Yao YG. Is there an antagonistic pleiotropic effect of a LRRK2 mutation on leprosy and Parkinson’ s disease? Proc Natl Acad Sci U S A. 2020.

34. Raza A, Raina J, Sahu SK, Wadhwa P. Genetic mutations in kinases: a comprehensive review on marketed inhibitors and unexplored targets in Parkinson’s disease. Neurol Sci. 2025.

35. Dong-Chen X, Yong C, Yang X, Chen-Yu S, Li-Hua P. Signaling pathways in Parkinson’s disease: molecular mechanisms and therapeutic interventions. Signal Transduct Target Ther. 2023;8(1):73.

36. Sauve V, Stefan E, Croteau N, Goiran T, Fakih R, Bansal N, et al. Activation of parkin by a molecular glue. Nature communications. 2024;15(1):7707.

37. Chin RM, Rakhit R, Ditsworth D, Wang C, Bartholomeus J, Liu S, et al. Pharmacological PINK1 activation ameliorates Pathology in Parkinson’s Disease models. bioRxiv. 2023:2023.02.14.528378.

38. Lambourne OA, Bell S, Wilhelm LP, Yarbrough EB, Holly GG, Russell OM, et al. PINK1-Dependent Mitophagy Inhibits Elevated Ubiquitin Phosphorylation Caused by Mitochondrial Damage. Journal of medicinal chemistry. 2023;66(11):7645–56.

39. Clark EH, Vazquez de la Torre A, Hoshikawa T, Briston T. Targeting mitophagy in Parkinson’s disease. J Biol Chem. 2021;296:100209.

40. Silvian LF. PINK1/Parkin Pathway Activation for Mitochondrial Quality Control - Which Is the Best Molecular Target for Therapy? Front Aging Neurosci. 2022;14:890823.

41. Chakraborty J, Ziviani E. Deubiquitinating Enzymes in Parkinson’s Disease. Front Physiol. 2020;11:535.

42. Durcan TM, Kontogiannea M, Bedard N, Wing SS, Fon EA. Ataxin-3 deubiquitination is coupled to Parkin ubiquitination via E2 ubiquitin-conjugating enzyme. J Biol Chem. 2012;287(1):531–41.

